# Evidence-based pandemic preparedness: an infrastructure for population-scale genome-based based testing for COVID-19

**DOI:** 10.1101/2021.06.10.21258526

**Authors:** Hans Lehrach, Jon Curtis, Bodo Lange, Lesley A Ogilvie, Richard Gauss, Christoph Steininger, Erhard Scholz, Matthias Kreck

## Abstract

Our lives (and deaths) have been dominated for more than a year by COVID-19, a pandemic that has caused hundreds of millions of disease cases, millions of deaths, trillions in economic costs, and major restrictions on our freedom. We argue that much of this could have been avoided by repeated and systematic population-scale PCR-based testing and targeted quarantine. We describe key elements of the current implementations of such a system and demonstrate (with Germany as an example), that this strategy could have suppressed the pandemic within weeks, eliminating the vast majority of its overall impact in terms of deaths, economic costs and restrictions. It can, however, still play a major role in further reducing the worldwide impact of the current phase of the pandemic, and remain as a key protection against similar dangers in the future.

---

To combat the spread of COVID-19, most countries have relied on social distancing, lockdown, limited testing and contact tracing procedures, as well as crash programs to develop vaccines, which are now increasingly contributing to the fight against the pandemic. As a complement to these two basic strategies, we and others proposed to use an alternative method, repeated population-wide tests^1-3^, to identify (and quarantine) the infected and break infection chains. This strategy has the potential to reduce human suffering, economic costs and restrictions on personal freedom to the inevitable minimum, since quarantine would be limited to a short time for a small fraction of the population and could be much more effective than inherently leaky lockdowns.

Through this, we could control or even eliminate the pandemic safely, quickly and comparatively inexpensively regionally, nationally and potentially worldwide. In spite of these likely advantages, and an early successful test of the strategy in Vò, an Italian city with 3300 inhabitants^4^, few governments seem to have seriously considered this as an alternative and, with the exception of Austria (‘Alles gurgelt’)^5^, none seem to have supported the establishment of the required infrastructure. Pilots of population-wide tests have been carried out, but have often been restricted to a single test cycle and/or relied on antigen-based rapid tests, which are inherently limited in their ability to stop new infections.

This lack of consideration for one of the most potentially powerful strategies in our arsenal against COVID-19, might have been due to a range of concerns regarding feasibility, from establishing the necessary low cost, highly sensitive test strategies to population-wide logistics and the cost-benefit ratio of such an approach.

To counteract these arguments, we describe here successful implementation of the two main elements needed: sensitive, scalable and cost-effective PCR based tests and a logistic infrastructure for population-wide tests. Using mathematical modelling, the enormous impact this strategy could have had on the pandemic in Germany is demonstrated; a country still dealing with the brutal consequences of COVID-19, with over 85,000 deaths, hundreds of billions of euros in economic costs and continuing restrictions on social freedoms for all. The overwhelming majority of this impact, including close to 90% of deaths, was caused by the second and third waves of the pandemic, and could therefore have been potentially avoided by PCR based mass testing.

## Results

### An affordable, scalable and highly sensitive virus-genome-based testing approach

A key component of the proposed strategy has been the development of highly-scalable, cost effective techniques to detect the viral genome with high sensitivity and specificity before infected persons becomes infectious themselves, in millions and ultimately billions of samples. This basically rules out the use of antigen-based rapid tests, which typically miss the first three days of a ten-day-long infectious period^6^. Standard qPCR tests, with their ability to identify individual genome molecules would be sufficiently sensitive: a new infection can only take place after an eclipse phase, which ends sometime after detectable RNA concentrations have been reached^7^. They are, however, not sufficiently scalable (e.g. total PCR test capacity in Germany falls short by >2 orders of magnitude) and the cost would be prohibitive.

We have therefore developed a x10-x20 more efficient, highly sensitive virus-genome-based test, using PCR technologies developed by some of us as part of the Human Genome Project^8^, and used to genotype billions of samples over the last decades^3^. All equipment is commercially available. Since we use the same reactions as standard qPCR tests but read the result differently (endpoint measurement^3^), we achieve the same sensitivity and specificity in a much more scalable fashion and at much lower costs per sample (∼€1 per PCR test for very high throughput). See Figure 1 for overview of the test pipeline. The procedure is EU-wide CE approved in gargle- and smear-based versions and is already being used in Germany to test the employees of Unilever and other companies. The key advantage is that infected individuals can be identified days earlier than with the antigen-based rapid tests currently used by the vast majority of companies; a prerequisite to stopping the spread of the virus. The test is highly scalable: A single commercially available water bath PCR system with a capacity of 100x 384 PCR plates per run would be able to carry out >600,000 RT-PCR reactions per day, close to three times the entire PCR test capacity currently available in Germany. A potential limitation is still the supply of test tubes and pipette tips (one per sample, far less than for standard qPCR tests). However, equipment to wash and reuse pipette tips is commercially available and approved for COVID-19^9,10^ and machines to reuse the sample tubes could be rapidly developed based on similar principles.

**Figure 1:**
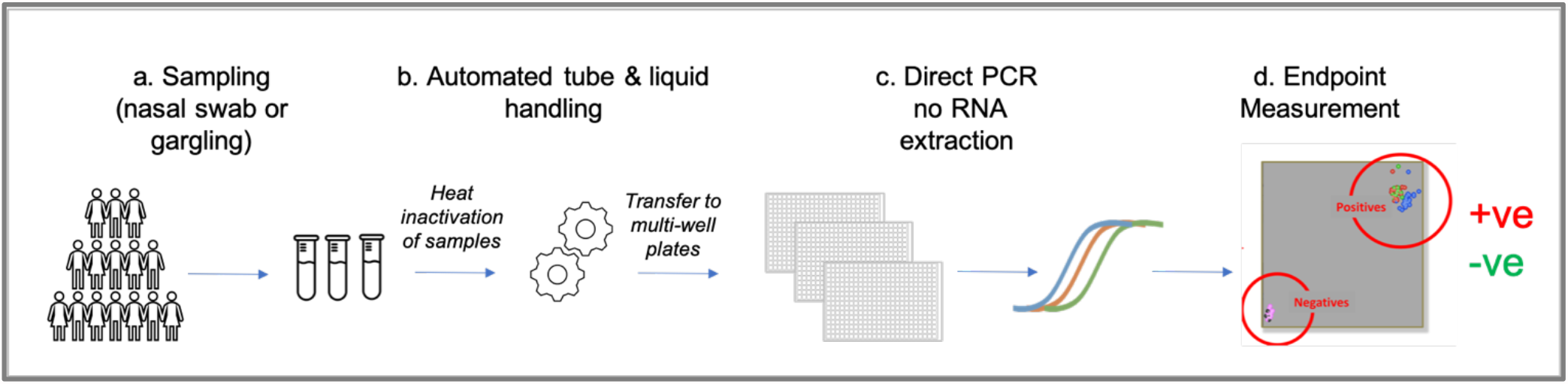
Schematic of the high throughput genome -based testing pipeline for SARS-CoV-2. (a) Sample tubes (barcoded) are brought to testing centres in 96-well carriers from collection points (e.g. see Figure 2) and prepared for high-throughput testing for SARS-CoV-2 (or other viruses). Samples are heat inactivated before automated transfer in 384-well format (b) and preparation for (c) direct RT-PCR without a requirement for RNA extractions and (d) endpoint measurement showing results of test (positive, negative).

The approach can also, in a slightly modified form, be easily used to identify sequence variants^11^. We will therefore be able to identify the most relevant groups of variants (e.g. UK, B.1.1.7; South Africa, B.1.351; Brazil, P.1, P.2; India B.1.617, B.1.617.1, B.1.617.2, B.1.617.3) in a second analysis cycle with a small number of additional tests on SARS-CoV-2 positive samples.

### Logistics: population-scale proof-of-concept

As a proof-of-concept, the government of Vienna, Austria, established a large, state-wide screening program which demonstrates the feasibility of such an approach (‘Alles gurgelt’)^5^. The aim of this program is the early interruption of infection chains, as well as tracing and prevention of spread of SARS CoV-2. For this purpose, a novel logistics concept was developed based on self-collected mouth wash samples. Self-collection of samples is done with support of a dedicated Web App, which guides the user through the procedure and validates their identity by verification of an identity card and video-surveillance of the procedure (www.lead-horizon.com), so that quality and reproducibility of sample-taking can be ensured.

The testing program is based on the promise that a reliable PCR test result is available for every inhabitant within a maximum of 24 hours after sample collection. All inhabitants were invited to participate in this program twice a week as regular testing makes sure that persons who have tested positive for COVID-19 can be quarantined and chains of infection interrupted early. Test kits for the validated self-collection of mouth-wash samples are distributed through local drug stores, which makes PCR tests available to all inhabitants within reach of a 5-minute walk. Following packaging in biohazard safe, sealed transport bags and cardboard packaging, samples are returned for transport to the laboratory at drug and grocery stores in Vienna.

Sampling devices are made accessible free of charge to all people living in Vienna, tourists and commuters. The successful program is currently being expanded in more pilot regions of Austria for further evaluation of a full, nation-wide coverage of the program. As an alternative, a similar kit based on nasal swabs, which might be easier to interface with the high throughput testing pipeline described here, has been developed at Alacris Theranostics. See Figure 2 for an outline of how the population-scale testing works in practice.

**Figure 2:**
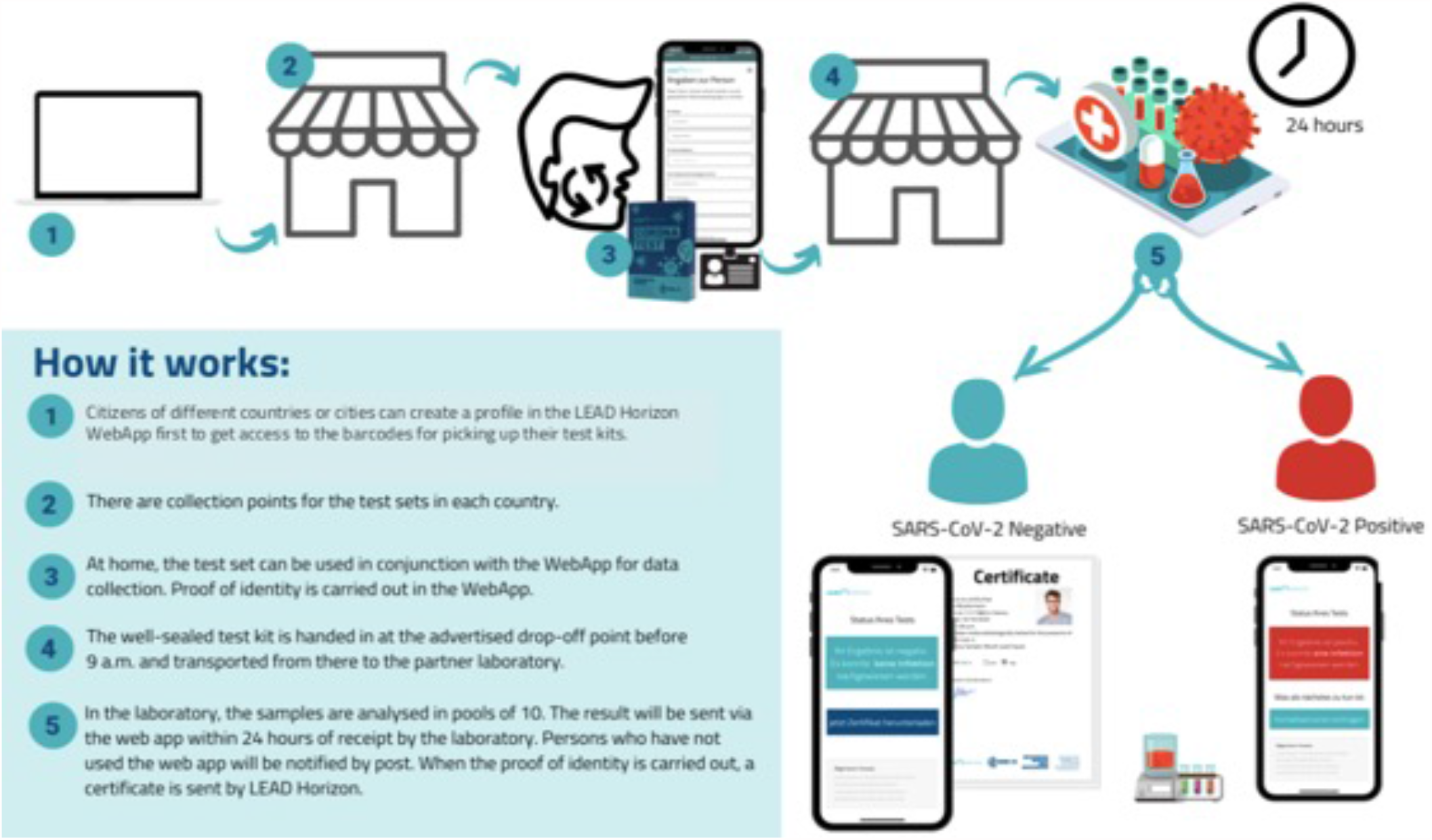
Population-wide testing logistics. An example of the decentralized test strategy used in Vienna.

### Modelling the impact of population-scale testing in Germany

To model the impact of repetitive population wide (or near-population-wide) testing, we show the effect this approach could have had on the development of the pandemic in Germany if population-wide tests were started last autumn, using a model based on the established Kermack-McKendrick theory, adjusted to COVID-19. The model and its validation are described in ref. ^12^. Besides reflecting changes of contact rates, the model allows to build in the effects of vaccinations and the developments of new mutants. To simplify the analysis, it is assumed that tests are 100% reliable and a certain percentage of the population is tested on a daily basis (see Kreck & Scholz^12^). Depending on how large this percentage is, one can predict what would have happened in Germany if such tests would have started last autumn (October 15^th^ 2020); a realistic scenario, since there would have been enough time to develop the required technology and infrastructure, if these developments had been supported by the government or private sources. In Figure 3, the numbers of daily new registered infected are displayed and corresponding R-values are shown, under the assumption that 60%, 80% or 100% of the population agreed to be tested on a daily basis using PCR and antigen-based tests, respectively.

**Figure 3:**
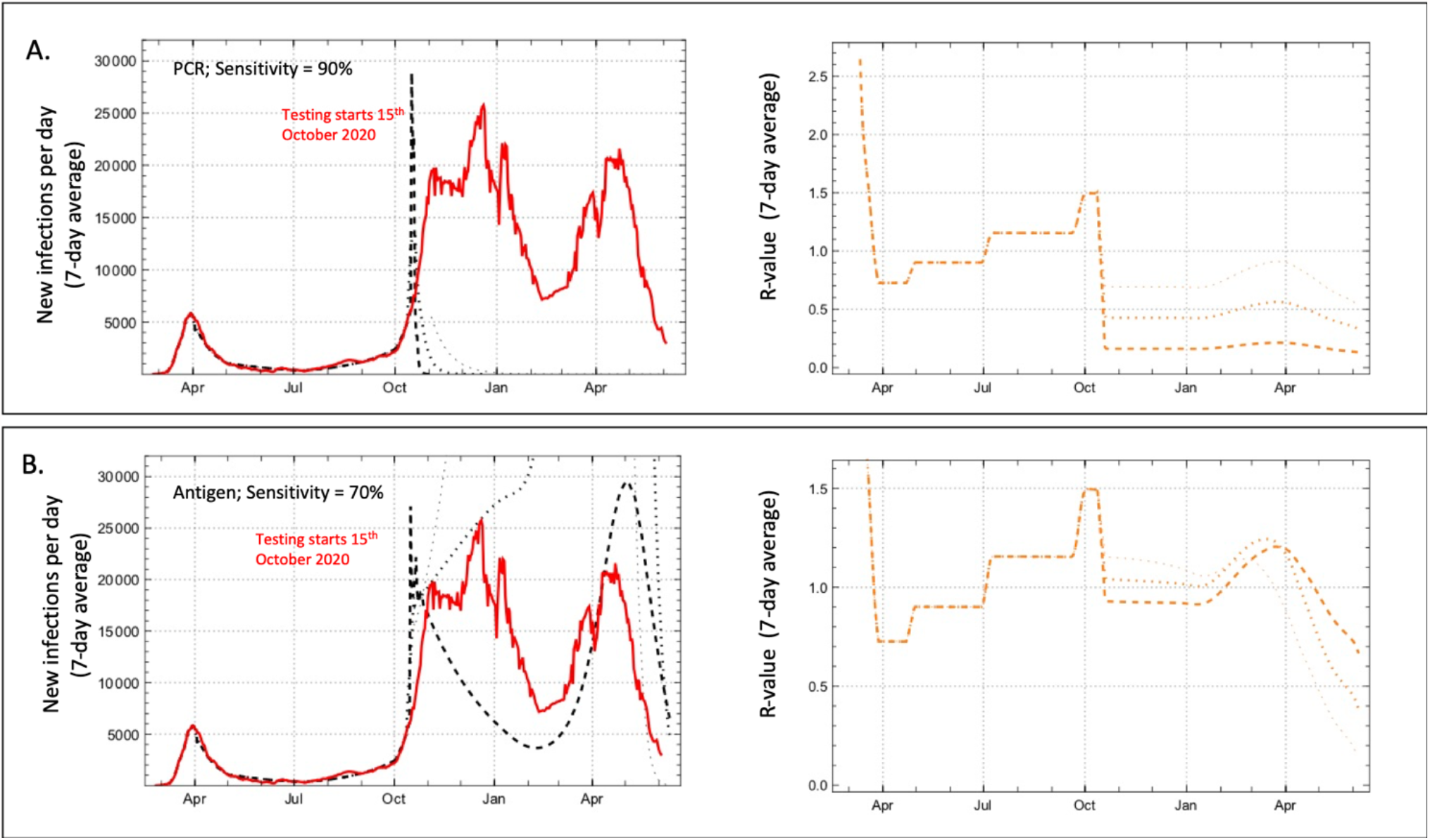
Modelling the impact of population-scale testing in Germany. Graphs show comparison of development of registered daily new infections (7-day averages) of John Hopkins University data for Germany (red line) with expectation and corresponding R-values under a test regime with daily testing using PCR (A) and (B) antigen-based tests, assuming participation rates of 60% (········), 80% (········) and 100% (———) of the population, and no further contact restrictions than those already operative in late October 2020 (black dashed lines). Left - new infections per day; Right - R-values.

### The scenarios assume the following conditions

- PCR tests are effective the day people get infectious, with a sensitivity of 90%. False positives are ignored for the model calculation (corresponding to a hypothetical specificity of 100%).
- Antigen-based tests are effective four days after a person is infectious and have a sensitivity of 70%.
- There are no extra restrictions compared to October 2020, i.e. the assumption is that the contact rates are more or less unchanged since this time. In reality, in the autumn and winter further strong restrictions were imposed in Germany; it is shown here that such restrictions would not have been necessary if the test regime had started.
- Vaccinations started in Germany from January 2021 onwards (as in reality).
- The UK virus variant (B.1.1.7), which is assumed to be 50% more transmissible, began to act from January 2021 onwards, dominating in early April 2021.

If the sensitivity falls moderately below 90% (the values assumed for the model calculation) the picture does not change qualitatively. A specificity of 100% may be assumed for the model, because the specificity of PCR reactions is very high (two specific primers, one specific probe), and can be further increased by, e.g. running a second cycle of analysis to identify the specific variant. Technical errors (cross-contamination of samples etc.) can be excluded by appropriate controls. In the calculation, effective enforcement of quarantine rules is assumed. Low levels of non-compliance would effectively decrease the overall participation rate, without major effect on the result. The model calculations therefore demonstrate convincingly the effectiveness of mass tests under only moderate contact restrictions like those in Germany in early October 2020. Even the rise of the new UK virus variant in early 2021 could have been kept under control by mass testing using highly sensitive PCR based tests, suppressing the R factor from 1.5 before the onset of testing to very low levels for a high degree of participation. Even low levels of participation (60%) are sufficient to reach R values far below 1, and therefore to suppress the pandemic. This is not the case at all if we model the effect of the antigen rapid tests, which are less sensitive, but also, more importantly, only become positive after onset of symptoms fairly late in the infectious phase. In this case, only 100% participation in daily testing would have reduced the R factor slightly below 1, only to return to values above 1, when the UK variant arrived. The drop in new infections shown in Figure 3 for the 80% and 60% scenarios would have been due to ‘herd immunity’ after very high levels of infection.

## Discussion

### Evidence-based pandemic preparedness?

We have been able to prove that a proposal to eliminate COVID-19 by systematic population-wide screens was very feasible. The simulations conducted comparing PCR and antigen-based tests, indicate that population-wide PCR-based tests even at fairly low levels of participation during the second wave (October 2020) could have resulted in a rapid drop in numbers of infections, to levels manageable by contact tracing. In contrast, even daily testing of the entire population with antigen rapid tests could have only achieved a modest and temporary reduction in the number of infected, followed by much higher numbers of infections later on. Given the demonstrated feasibility and cost-effectiveness of this approach, it is hard to understand why this possibility was not explored further earlier in the pandemic. In view of the enormous impact this approach could have had on the course of the pandemic, this raises the question if the mechanisms governments use to evaluate potential solutions to problems of such enormous importance, should not be to more open and science-driven.

The pandemic is, however, far from over. Roll-out of vaccinations in many areas of the world has been much slower than originally anticipated (e.g. in India) and new variants threaten to undo the progress made. Indeed, recent reports seem to indicate that the AstraZeneca vaccine provides limited protection against the South African and Brazilian variants of the virus^13^. We should therefore guard against the possibility that new variants could also break the immunity generated by the other vaccines.

As a complement to the effort on vaccination, we should therefore urgently set up (and use) an infrastructure for virus genome-based mass tests to protect the population from increasingly aggressive SARS-CoV-2 variants. Such an infrastructure would also be able to respond to any new, similar threats (essentially all pathogens have genomes) within a few weeks rather than the year(s) new vaccine development is likely to take. We have the tools. We just have to be ready to use them.

## Methods

### High-throughput PCR testing

High-throughput PCR based tests are performed on either nasal swabs or gargle samples, collected, if possible in kits, useful for direct testing and eliminating the need for RNA purification. Samples are received in triple coded screwcap tubes in SBS carriers with 96 positions (including controls). After virus inactivation (as part of transport medium in the case of nasal swabs, by heat treatment for 30 minutes at 65 °C for gargle samples), the bottom barcodes are read (FluidX Perception HD Whole Rack Reader), the tubes are decapped using an 8 or 96 position decapper (e.g. FluidX XDC-96 Automatic 96-Format Tube Rack Decapper/Capper), and 4 µl (10 µl reactions) or 8 µl (20 µl reactions) are transferred (e.g. Biomek i7 Automated Workstation) using a 96 pipetting head into 384-well PCR plates already containing the appropriate master and assay mixes.

Plates are heat-sealed (e.g. PX1 PCR Plate sealer), and the contents are amplified by RT-PCR reactions using either standard high-throughput PCR machines (e.g. DNA Engine Tetrade 2 Thermal Cycler) or, for very high throughput, hydrocyclers (e.g. KBiosystems DT-24 Thermal Cycling PCR System). Plates are then read on a fluorescent plate reader (e.g. CLARIOstar Plus Plate Reader (BMG)) and scored automatically. Individuals are notified of the test results by SMS or email. In the case of positive tests, the local heath authority is also informed. Throughput is mostly limited by the RT-PCR capacity, and can reach >600,000 samples per day for a single 100x 384 well plate hydrocycler.

### Mathematical Modelling

The methods used to construct the model are described in ref. 12. The model is based on the established Kermack-McKendrick theory, adjusted to COVID-19 and implemented as Mathematica 12 notebooks. In the model, the effect of population-wide testing of specific fractions of the population was superimposed on the effects of the series of social distancing/lockdown measures taken or the social distancing/lockdown measures in force at the assumed beginning of the tests (15^th^ of October). In predicting the later development of the pandemic, both the effects of new variants (UK variant from January 2021) and vaccination (also from the beginning of the current year) are taken into account.

The three notebooks used in this work are available for download through http://www2.math.uni-wupper-tal.de/~scholz/model-epidem.html.

## Data Availability

The three notebooks used in this work are available for download through http://www2.math.uni-wuppertal.de/~scholz/model-epidem.html

## Acknowledgements

The authors would like to thank Prof. Karin Mölling and Prof. Ulrike Protzer for discussions on SARS-CoV-2 testing and Dr. Sebastian Meier-Ewert for many productive discussions on large scale-test strategies. In addition, we thank the team at Alacris Theranostics GmbH, in particular Dr. Andelé Conradie and Dr. Artur Muradyan, for their contributions to the test platform and constructive criticism and feedback on the manuscript.

